# Lockdown impact on age-specific contact patterns and behaviours in France

**DOI:** 10.1101/2020.10.07.20205104

**Authors:** Paolo Bosetti, Bich-Tram Huynh, Armiya Youssouf Abdou, Marie Sanchez, Catherine Eisenhauer, Noémie Courtejoie, Jérôme Accardo, Henrik Salje, Didier Guillemot, Mathieu Moslonka-Lefebvre, Pierre-Yves Boëlle, Guillaume Béraud, Simon Cauchemez, Lulla Opatowski

**Author notes:** contributed equally (Paolo Bosetti, Simon Cauchemez, Lulla Opatowski). Corresponding author: Paolo Bosetti.

## Abstract

In the first trimester 2020, a significant number of countries implemented general lockdowns of their populations to contain the quickly expanding SARS-CoV-2 epidemic and avoid major saturation of health care capacity. Understanding how these unprecedented measures impacted population behaviour and contact patterns is key to predict more accurately the health, social and economic impacts of such extreme actions if they were to be applied to future outbreaks. We set up an online survey to measure how the lockdown affected social contact patterns in France, and collected information from 42,036 participants aged 18 years and over between April 10 and April 28, 2020. Among the participants who normally worked outside home prior to the lockdown (72% of the survey population), 68% reported that they had moved to working from home and 17% reported being unemployed during the lockdown.

Only 2% of participants used public transport during lockdown, as opposed to 37% before it. Participants reported increased frequency of washing hands, switch in greeting behaviour, but generally limited use of masks outside home. 138,934 contacts were reported, with an average 3.3 contacts per individual per day (1.7 for individuals aged >65 years old compared to 3.6 for younger age-groups). This represented a 70% reduction compared with previous surveys, consistent with reductions in transmission rates measured during the lockdown. Contacts in workplaces, shops, and transports on the previous day were respectively reported in only 11%, 31% and 0.5% of the participants. For those who maintained a professional activity outside home, the frequency of contacts at work dropped by 79%. This study shows that the lockdown dramatically affected population’s behavior, work, risk perception and contact patterns. Both frequency and heterogeneity of contacts were affected, impacting potential important features of virus dissemination. Such surveys are essential to evaluate more accurately the impact of past or future lockdowns and anticipate epidemic dynamics in these conditions.

## Introduction

Following its discovery in Wuhan province in China in December 2019, the virus SARS-CoV-2 has quickly spread around the world. On March 17th 2020, the French government implemented a lockdown of its population to attempt to contain the quickly expanding epidemic and avoid a saturation of health care capacity. This lockdown consisted of a set of interventions including school closures (schools remained open for children of healthcare and other essential workers), closure of universities, restaurants, non-essential shops and most workplaces. A move to working from home was promoted where possible. Outdoor physical exercise was restricted to no more than 1 hour a day and no further than 1km from home. To help ensure compliance, individuals had to fill in a form each time they got out, stating the purpose of their journey out of a limited list of possible options.

Understanding how these unprecedented measures impacted population behaviour and contact patterns is important to better describe SARS-COV-2 transmission dynamics during the lockdown period. On the one hand what truly matters for transmission are the social, cultural and behavioural responses to disease, public communications (through official or media channels) or strict measures (1,2). Mathematical models used for outbreak predictions usually rely on empirical data describing the rate at which individuals mix with each other according to age. Although such data is available from surveys performed in multiple countries in a non epidemic situation(3)(4)(5)(6), depicting contact patterns during lockdown is crucial.

## Methods

### Survey

We set up SocialCov, an online survey to record social contact patterns and behaviours within the French population. The survey was advertised on social networks (including posts on Facebook and Twitter), messaging platforms (WhatsApp), emails, and the Institut Pasteur website. All users over 18 years old were invited to fill in the questionnaire. Information collected included socio demographic information : age, sex, usual place of residence, number and age of usual household members and employment characteristics, then whether changes occured in the place of residence, household composition and employment status during the lockdown.

Participants also reported information about their contacts and their contacts’ characteristics, including the age of the individual and the place of contact (work, transportation, medical appointment, sport practices, shopping, assistance to a person in need) on the previous day. A contact was defined as either a physical contact (e.g. a kiss or a handshake), or a close contact (e.g. face to face conversation at less than 1 meter). Data regarding childcare arrangements, lifestyle habits (shopping frequency, mode of transport, etc), and individual protective behaviours (hand washing, mask wearing, etc) were also collected. Participants were asked to provide this information for the previous day, which was during lockdown, and on a typical day before the lockdown.

Data were collected in accordance with the regulation in force in France for the protection and security of personal data. Aggregated data is accessible online (link here). The complete dataset can be provided on demand, subject to prior determination of the terms and conditions of the request and in respect for the compliance with the applicable regulation.

### Statistical methods

We analysed data collected between April 10 and April 28, 2020. We grouped study participants and contacts into the following age groups: 18-20, 21-25, 26-30, 31-35, 36-40, 41-45, 46-50, 51-55, 56-60, 61-65, 66-70, 71-75, 76-80, 81-85, 86-90, >90 and computed descriptive statistics for the responses related to age, household, location, activity, and individual preventative measures. We present either the distributions of numbers and percentages or means and standard deviations as appropriate.

#### Contact matrices

We computed the average number of contacts per person per day stratified by age group, gender, work activity and place of contact and built contact matrices. For home matrices, all individuals of a household were considered as close contacts. For other locations (work, shops, public transports), close contacts and age of contacts were computed by summing those reported by the participant in the corresponding setting. Outlier reports were defined as participants reporting more than 100 contacts per day or more than 30 contacts per day outside the work and household location. These outlier reports were removed from the analysis. A global matrix, reporting the average total number of contacts per day across ages, was also computed. In this matrix, for each age group, the average number contacts was reweighted to account for gender and professional activity distributions in the French population during lockdown.

### Other source of data used for population correction

In order to limit bias due to the study design and potential lack of representativity of the study population, estimates were corrected using metrics from the following data sources.

#### INSEE

Demographic data of the French population reported by the French National Institute for statistics and economic studies (INSEE) was used to compute weighted estimates and reproduce the age and sex distribution in the French population of those aged >18 year old(7). Weighting was achieved for all estimates related to professional situation and lockdown associated behaviours. No weighting was done to compute descriptive statistics related to the survey population.

#### Covid 19 Barometer

*Covid 19 Barometer* is a separate survey from the non-profit organisation DataCovid, which was carried out during the same period. In this survey, weekly polls were administered on the internet by the company Ipsos with samples of 5,000 people representative of the French metropolitan population aged 18 and over established by the quota method (see Supplementary Material section B for more details). Estimated professional activity distributions by gender were used as a reference measure to compute corrected estimates of the global matrix.

In the following, when presenting unweighted data, we use the term participant, when weighted data, we use the term French population.

### Comparison of contacts with pre-lockdown situation

We compare the frequency of contacts during the lockdown with a non-epidemic period, using direct reports of participants from the SocialCov survey (the part related to their contacts on a day preceding lockdown) and data from a previous survey in France, the COMES-F (5).

#### Comparison with SocialCov pre-lockdown survey

Participants involved in the SocialCov survey were also asked to fill an additional pre-lockdown survey and to depict their contact at the workplace for a day preceding the national lockdown. A total of 35982 participants completed this second questionnaire. We used the full dataset to compute the distribution of the number of contacts. We considered only the participants that declared to work outside home in the pre-lockdown period and during lockdown (3186 participants) to assess the individual change between the number of contacts before and under the lockdown.

#### COMES-F survey

The COMES-F study was the first French large-scale population survey performed by Ipsos in 2012 (5). The aim of the study was to describe the mixing pattern by age in the French population. In total 2033 participants reported their contacts over 2 days, one during the week day and one during weekends. To match and compare the two datasets, individual contacts in SocialCov were censored at 40 daily contacts to comply with COMES-F constraints. Also, only contacts during weekdays were considered for the COMES-F survey. Censoring all contacts >40 generated a reduction of 0.2 in the average number of contacts in our survey, from 3.3 without censoring to 3.1 with censoring.

## Results

### Population description

42,036 participants completed the full questionnaire between April 10 and April 28 (42,130 in total and 94 were classified as abnormal reports), including 28,796 females and 13,240 males from across France (Figure S1A), with an average of 4.4 participants per 10,000 inhabitants (range from 0.89 per 10,000 in Haute Corse to 29.5 per 10,000 inhabitants in Paris). The region of Paris Île de France, and departments of Doubs, Aude, Haute Garonne, and Herault were over-represented, with >10 participants per 10,000 inhabitants (Figure S1B, Table S1). Half of the participants were <45 years old and 13% were >65 years. The number of household contacts was the highest for participants aged 18-20, subsequently dropping for those in their 20s and increasing again to a second peak for individuals in their late 40s (Figure S1C). Participants >60 years old only had about 1 household contact on average. 37% of participants declared being locked down with children (<18 years old) in the household. Only 2.5% of participants aged >60 years old declared a contact aged <18 years old at home, compared with an average of 77% for those aged 41-45 (Figure S1E).

### A major drop in contact frequency even for the more connected individuals

In total, 138,934 contacts were reported, representing an average number of contacts of 3.30 per day (median = 2 range [0-99] daily contacts) per participant after removing outlier reports. Individuals aged >65 years old reported on average 1.67 contact per day compared with 3.55 contacts per day for those aged <65 years old (Figure 1A). These estimates are 70% (range: 61%-76%) lower than those measured in the COMES-F survey performed in France in 2012 (Figure 1A) (5).

**Figure 1.**
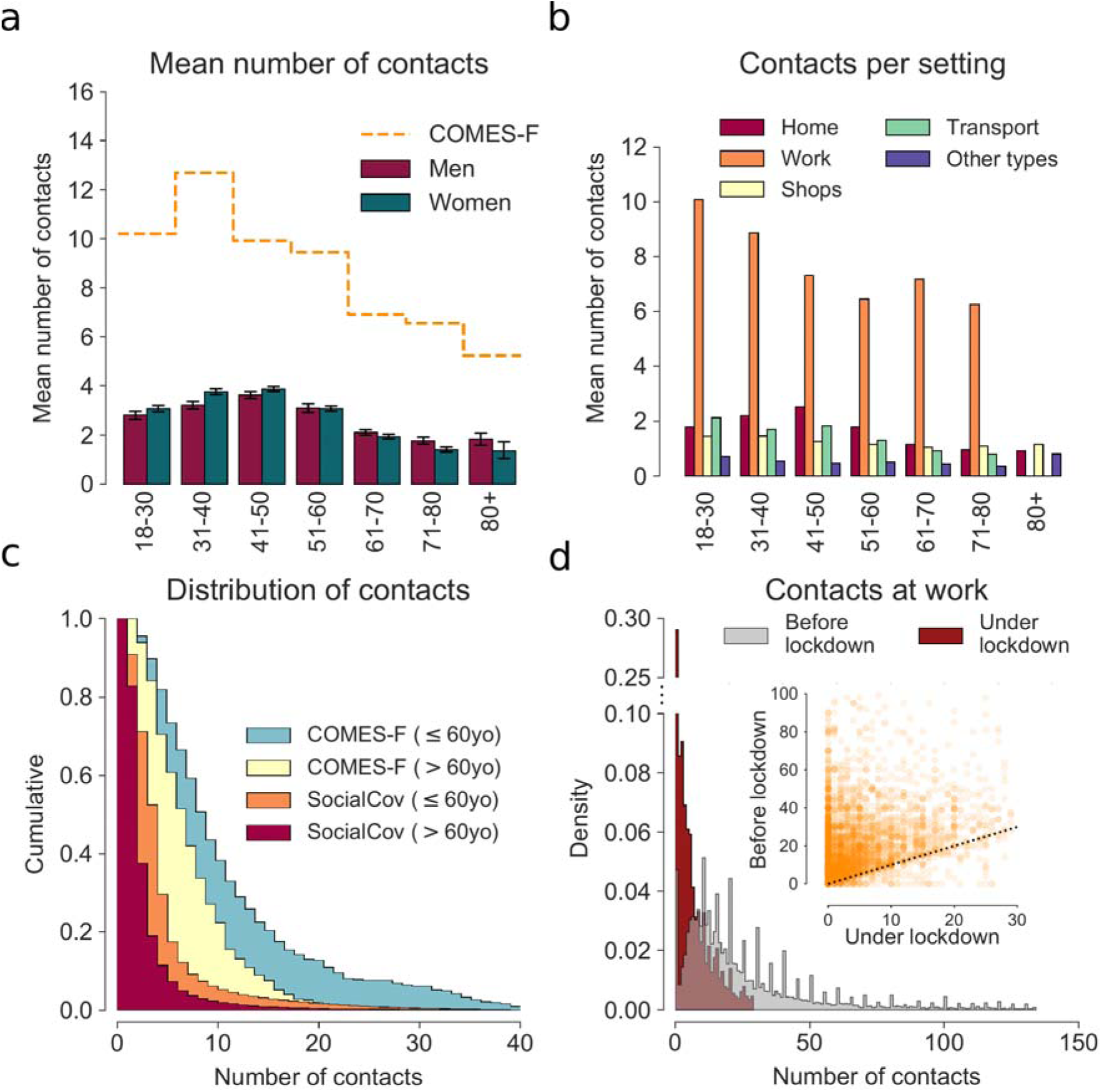
Number of contacts by age-group and location during lockdown. a) Average number of contacts depending on the age of participant. For each age-group (x-axis), the bars represent the mean and 95%CI of the number of contacts reported by participants, whatever the location. Orange line represents the mean number of contacts for the COMES-F study. In order to compare the two dataset, individuals with more than 40 contacts per day were not considered. b) Average number of contacts depending on the location. For each age-group (x-axis), bars represent the mean number of contacts for each setting reported by participants. c) Light blue and yellow curves show the cumulative distributions of the number of contacts for participants in the COMES-F dataset for ages between 18 and 60 years old and more than 60 years old, respectively. Orange and red curves represent the cumulative distributions of the number of contacts for participants in the SocialCov study for ages between 18 and 60 years old and more than 60 years old, respectively. In order to compare the two dataset, individuals with more than 40 contacts per day were not considered. d) Distribution of the number of contacts at work before lockdown (grey), and under lockdown (red) only using observations up to the 95 percentile. Inset shows the change in the number of contacts before and under lockdown at the workplace for participants working outside home during the lockdown (orange).

Most participants declared working from home during lockdown (N=22,327) or being unemployed or retired (N=15,142). 4,567 (11%) participants reported contacts at work; 12,967 (31%) contacts in shops; and only 203 (0.5%) contacts in public transports. Finally, 12,325 (29%) participants reported contacts in other places, which included medical appointments, physical activity and visits/assistance to relatives in need but with a much lower intensity than in other settings. The average number of daily contacts in each setting is shown on Figure 1B. The density of contacts was the highest at work, with the average number of contacts ranging 6-10 depending on the age group. It was much lower in other settings, with between 0 and 2 contacts per day on average.

Critically, the proportion of individuals with a higher number of contacts markedly decreased during the lockdown (Figure 1C). Specifically, only 4.9% of the participants aged 18-60 years old reported more than 10 contacts per day compared to >35% in the COMES-F study (5). This percentage decreased to 1.7% among the elderly in our sample while it was 18.8% in the COMES-F study. Similarly, among those who maintained their professional activity outside home, the intensity of contacts at the workplace was reduced by 79% between the week preceding the lockdown and the lockdown period, with a distribution shifting towards lower values, approaching zero in the majority of cases during the lockdown (Figure 1D).

In addition to contacts of study participants, some information on children’s contacts within and outside the household were available through participants’ reports. All in all, during lockdown, the vast majority of children’s contacts occurred in the household. Indeed, only 5% of participants with children had their children cared for outside home, with school or nursery attendance reported only by 2% of participants. Only 3.4% (N=508) of the children for which contact information was reported had contacts with children that were not household members (Figure S6), with an average of 2.5 contacts per day. This represents a drastic decrease compared with previous surveys (5).

The total number of daily reported contacts also varied between geographic areas. In departments with >500 participants, the average number of daily contacts varied between 2.7 (95%CI 2.4-3.0) in Alpes-Maritimes, and 4.4 (95%CI 3.8-5.0) in Seine-Maritime. Participants from geographic areas with higher densities generally reported higher frequencies of contacts outside home and work (Figure S7, r^2^= 0.48).

### Lockdown affected population behavior

Consistent with studies of mobility (8, 9), we identify a pattern of migrations from big cities to other confinement locations at the time when the lockdown was implemented. In our survey, Paris was by far the area that exhibited the highest level of migration, with about 20% of participants from Paris declaring being in another department during the lockdown (Figure 2A), predominantly in the West and South East of France (Figure 2B). Of interest, participants from Paris who declared being in a different geographic area, reported a lower frequency of contacts outside home than the ones who stayed in Paris (Figure S8).

**Figure 2.**
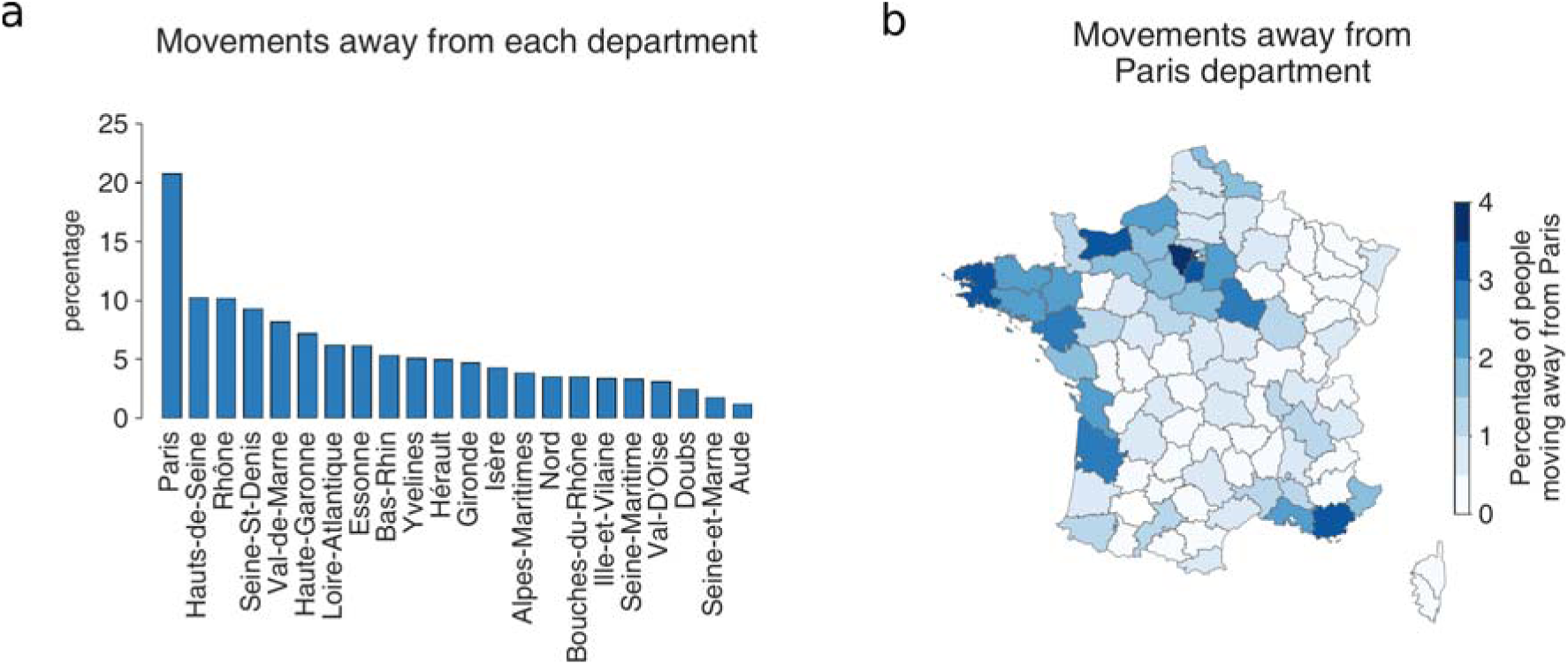
Spatial dispersion during lockdown in France. A. percentage of the respondents for which department of stay during lockdown differs from their home department. Only departments for which >500 answers are represented on the figure. B. Maps of department residencies during lockdown for individuals declaring 75 (Paris) as their home department.

We estimate that <2% of the French population used public transport during the lockdown, compared to 39% before the lockdown (Table S2). Shopping frequency also dropped in the population. Approximately 60% of the population used to go shopping >2 times a week in normal times, whereas only 14% went under the lockdown. Interestingly, 19% did not visit any shops at all in the preceding week (Figure 3A).

**Figure 3.**
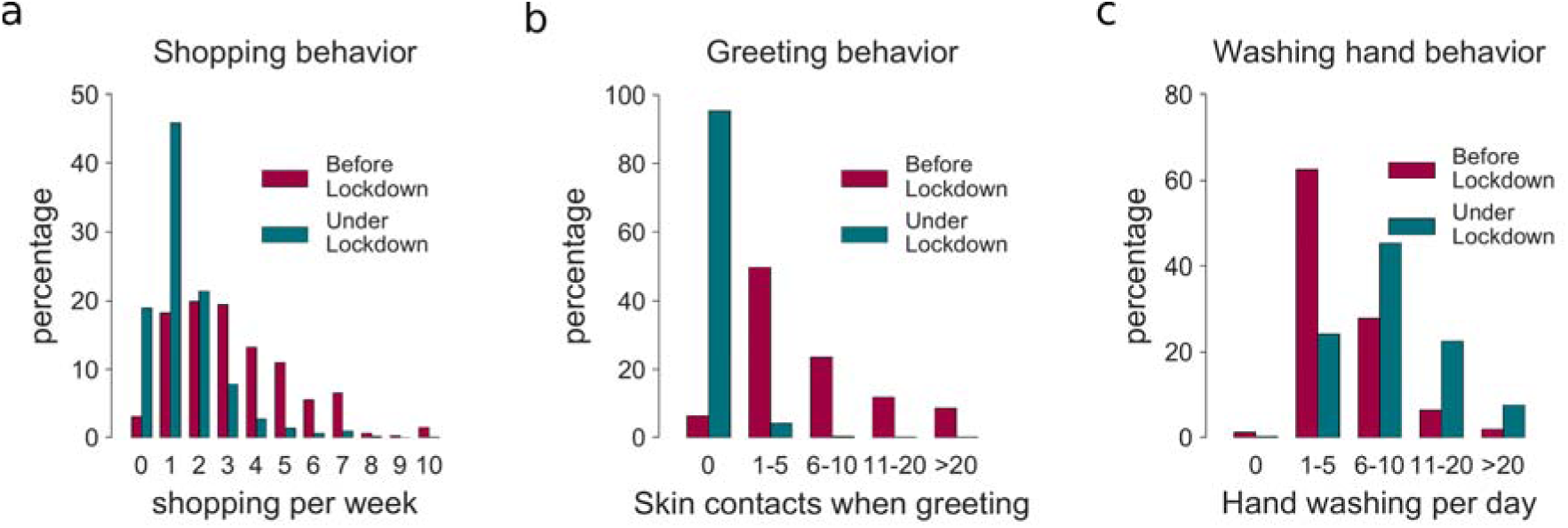
Lockdown associated behaviours. a) Change in shopping frequency. Distributions of shopping frequency (number of times individuals shop in a week) before (purple) and during (blue) lockdown. b) Frequency of hand hygiene before and during lockdown. Distributions of number of reported contact-associated to greeting in a typical day before (purple) and during (blue) lockdown. c) Change in greeting behaviour. Distributions of reported frequencies of hand hand washing in a typical day before (purple) and during (blue) lockdown. The percentages are weighted to account for the French population distribution (age and sex) reported by INSEE in 2019(10).

The outbreak and lockdown strongly affected risk perception and health-associated behaviours in the population. >60% of the population declared that their perception of the risk associated with the outbreak had changed following the start of the lockdown (Table S2). Only 1.5% declared perceiving no risk associated with the outbreak and more than 80% of the population reported a perceived risk for themselves and their relatives (Table S2). In terms of barrier measures, the percentage of the French population washing their hands more than 6 times a day increased from 36% prior to the lockdown to >75% under the lockdown (Figure 3C). Greeting behaviour was also strongly modified, with an estimated 95% of the population not greeting anyone outside the household physically (kissing or shaking hands), compared with only ∼6% before the lockdown (Figure 3B). While 47% of the population declared not systematically using masks outside home, 57% would wear one at home if symptomatic (Table S2). Finally, 81% of the population declared that they would not physically attend a non urgent medical appointment: 48% would cancel the appointment and 32% would use telemedicine (Table S2).

### Lockdown-associated unemployment and telework

After correction for population demographics, we estimate that during the lockdown, 44.2% of the French population older than 18 years old were unemployed or retired (compared to 33.9% before the lockdown), 46.7% worked from home (compared to 3.7% before the lockdown) and 9.1% worked outside home (compared to 62.3% before the lockdown) (Figure S2A-B). We estimate that among French people who normally work outside home, 68.4% switched to telework and 16.8% became unemployed or retired (Figure S2C).

### Age-stratified matrices show little cross-generation mixing of the elderly

Using participants’ reports of their contacts across age-groups, we built age-stratified contact matrices for each setting. The resulting home matrix is highly assortative by age group with a strong interaction term between parents and children (Figure 4A). In contrast, the contacts at work or in shops had little age structure (Figure 4 B-C), with an average number of daily contacts of 7.9 at work and 1.3 in shops for those who reported such contacts the previous day. The average number of contacts in public transport was low (computed based on 203 reporting participants only) with an average of 1.7 contacts per day and representing a very small fraction of the total number of contacts within the population (Figure 4E). Finally, the weighted global matrix depicts a higher density of within age group contacts and between-generation family-like contacts, consistent with previous findings. Of interest, individuals aged >65 years old had very limited contacts with the youngest age groups (<20) and had on average 50% less daily contacts than age groups 30-55 (Figure 1 and 4F).

**Figure 4.**
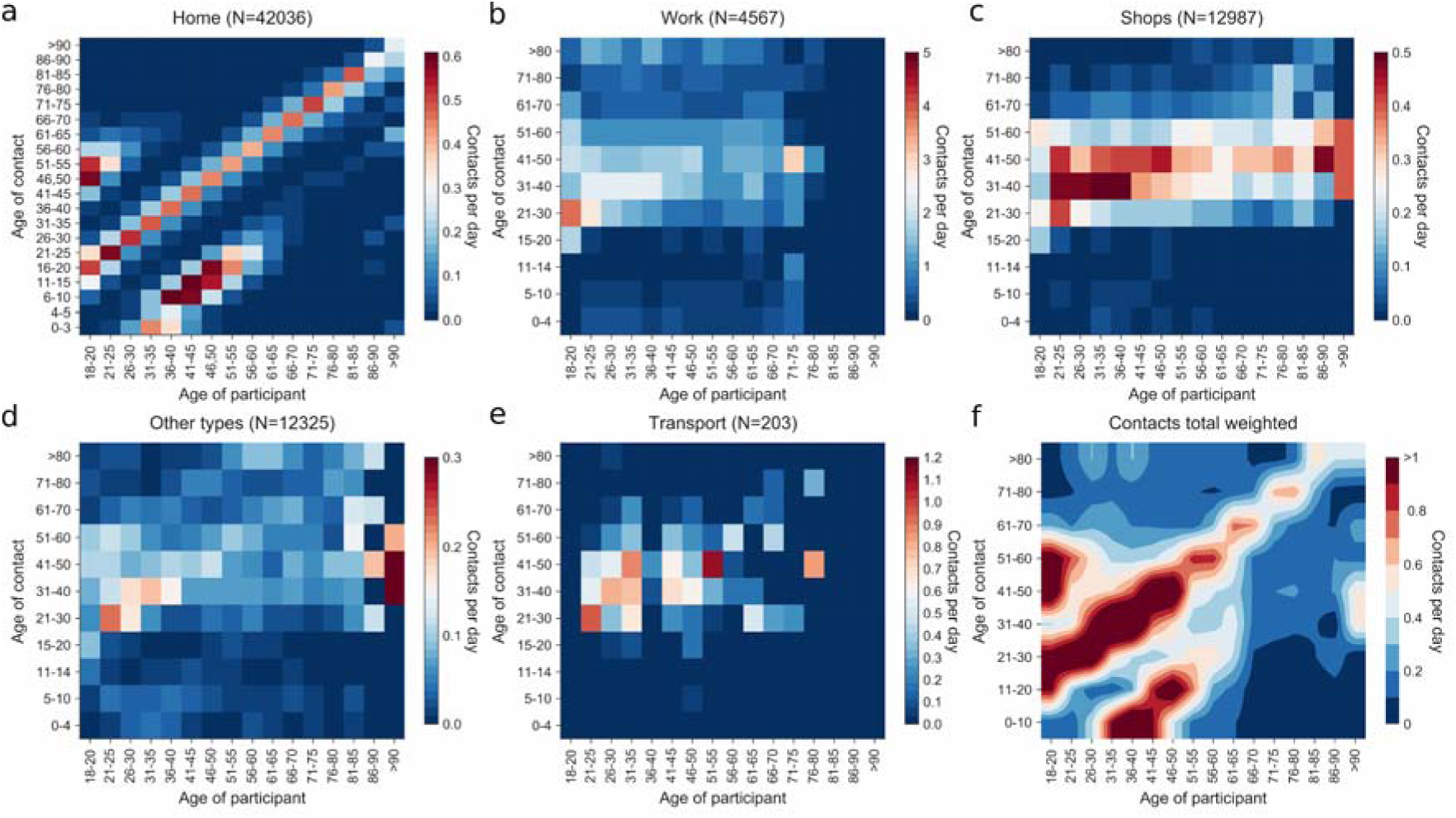
Contact matrices during lockdown in France. Frequency of individual close contacts at less than 1 meter per day according to age of participant (x-axis) and age of the contact (y-axis). A. Contacts occurring at home. B. Contacts reported at the workplace. C. Contacts reported in shops. D. Contacts on other locations. E. Contacts in public transports. F. All contacts weighted using the French population distribution (INSEE(10)) and professional activity distributions during lockdown (Covid 19 Barometre).

## Discussion

The SocialCov study reveals a strong impact of the lockdown on social patterns and behaviours in France, with over 40,000 responses from diverse ages and coming from all across France.

A drastic drop in contacts in all ages was observed, even in individuals who maintained a professional activity outside home. Notably, both the mean and the dispersion of contacts were affected. The tail of the distribution of the daily number of contacts was strongly diminished, with less than 5% of the participants reporting more than 10 contacts per day. Recent publications suggest a key role of superspreading events in the SARS-Cov2 outbreak(11). In general, decreasing the number of contacts in the most active part of the population might have led to fewer super spreading events and less virus diffusion.

A majority of our participants declared that they would either cancel a planned medical appointment or use telemedicine rather than attending in person. This reduction in care seekings contributed to a 51% reduction in consultations with specialized physicians (12), a 40% reduction in general practice consultations (French National health Insurance), and 48% reduction in emergency department admissions. This was also observed among individuals affected with chronic disease where 51% of the respondents declared they cancelled at least one medical appointment and 16% reported using telemedicine services (13). Estimating the indirect health burden associated with the lockdown will be critical in the coming months.

Only a small proportion of participants reported using protective masks outside home. These relatively small numbers may have been impacted by the limited availability of masks to the general population during the study period, when mask sales and distribution were restricted to healthcare professionals. It is expected that mask-associated behaviour changes with increasing availability of masks in the community and increased recommendation of using them.

A proportion of participants reported being locked down in a different department than their home department (Figure 2). This was particularly significant for participants living in big cities such as Paris (20%) or Lyon (10%). Our results are consistent with the analysis of mobile phone data indicating that 11% of the registered residents and 23% of the total number of people present in Paris the night before the lockdown moved to another location (9). Political elections took place in France on the weekend preceding the implementation of the general lockdown which could have led some individuals from Paris (eg. student or other non-official residents) to travel back to their original region to vote. This phenomenon might also be related to pre-lockdown travel that was also observed in other countries such as China (from Wuhan to other regions), Italy (from the North to the South)(14), Spain (from Madrid to other regions)(15) where people moved to their second residences or hometowns to avoid being quarantined in the city prior to the implementation of lockdown.

The estimated decrease in contact intensity is consistent with estimates from other studies in Europe(16), in the UK, Jarvis and colleagues reported a 74% reduction in the average daily number of contacts, with an average 2.8 contacts per day from 1356 participants in the UK (17). Similar reductions were observed in China, from 636 study participants in Wuhan, and 557 participants in Shanghai, where the average number of daily contacts was estimated at 2, representing a reduction of 7-8-fold during the COVID-19 social distancing period, with most interactions restricted to the household (18).

The data and results presented here should be considered in the light to the following limitations. First, participants were recruited online. As a consequence, the study population may not be a representative sample of the French population. First, as frequently observed in this type of studies, two third of survey participants were women. However, the work situation (Figure S9) and contact matrices (Figure S10) did not differ significantly between males and females. In order to reproduce more accurately lockdown-associated behaviors in the French population, statistics were reweighted according to the age and sex demographics of that population. The distribution of household sizes in the weighted population (Figure S3) globally matched the one reported by INSEE in 2019 (19).

The survey population is probably also over-represented by employees, executives and generally individuals working at home. It also excluded populations less connected to social networks, smartphones or internet. For example, >65 years old individuals represented only 13% of our population while they account for 20% of the French population. We estimate that during the lockdown 53% of French people worked from home, an estimate substantially higher than estimates from other sources (Direction de l’animation de la recherche des études et des statistiques, DARES or the *Covid 19 Barometre*, which estimated respectively 25% and 15% of telework). In order to bypass this overrepresentation, the global contact matrix was computed by weighting according to gender and professional status (telework, unemployed, retired) estimated from the *Covid 19 Barometre*.

This article provides novel data on social patterns during a strict lockdown. Following lockdown relaxing and school and workplaces reopening, intermediate and progressive changes in behaviour and social patterns are expected to have happened. Monitoring contacts, risk perception and behaviours following the ease of the restrictions will be key to assess the dynamics of contact patterns over time. In low and middle income countries, very little data related to social mixing in the community are available. These data are all the more needed to analyse the spread of the virus and the impact of interventions in these countries where social distancing and barrier gestures are potentially more difficult to implement and health care capacity, particularly intensive care, is limited. The set up and extension of similar surveys in the coming weeks and months will be critical.

From this survey, we estimated that the lockdown in France led to major reductions of social mixing across all age-groups, compatible with the estimated important decrease in reproduction number (20). The resulting matrices and data can be directly plugged into mathematical models of human-human transmission of SARS-Cov2. They may also be useful to model and evaluate the impact of lockdowns in future epidemic waves of SARS-CoV-2 or in future pandemics. Monitoring how contact patterns and at-risk contacts evolve over time will be key to anticipate the dynamics of the virus in the coming weeks and months.

## Data Availability

The complete dataset can be provided on demand, subject to prior determination of the terms and conditions of the request and in respect for the compliance with the applicable regulation.

## Acknowledgement

We thank Frederic Gouin, the data protection officer and Nathalie Joly, clinical core at Institut Pasteur for their advice.

